# Postpartum Exercise Promotes Maternal-Infant Molecular Communication via Breast Milk Small Extracellular Vesicles

**DOI:** 10.64898/2026.05.14.26353181

**Authors:** Devanshi Gupta, Kyle J. Sevits, Katherine A. Klaus, Shiana S. Loona, Vidhi D. Parmar, Yohan Kim, Carrie J. Heppelmann, Robert G. Leija, Hawley E. Kunz, Fabrice Lucien, Linda M. Szymanski, Aoife M. Egan, Mark W. Pataky

## Abstract

Early-life nutrition profoundly influences long-term metabolic health, and breast milk not only provides nutrients but also conveys maternal signals shaping infant metabolic development. While postpartum exercise by lactating women benefits maternal health, its impact on milk-borne signaling remains largely undefined. Small extracellular vesicles (sEVs) in breast milk are key mediators of maternal–infant communication because of their selectively packaged bioactive cargo and resistance to infant digestive enzymes and acids, enabling delivery of their cargo to peripheral tissues. Here, we show that a single session of moderate-intensity postpartum aerobic exercise robustly increases human breast milk sEV concentration, which persists for multiple post-exercise milk collections. Exercise enriches breast milk with sEVs containing regulatory metabolic cargo (proteins, miRNAs, and metabolites), which translates into enhanced mitochondrial capacity in neonatal-stage cells. These findings implicate sEVs as an exercise-responsive signaling compartment in breast milk capable of connecting postpartum maternal physical activity to beneficial infant metabolic programming.

**Highlights:**

- Acute moderate-intensity exercise increases human breast milk sEV concentration
- The exercise-mediated sEV increase lasts for multiple subsequent milk expressions
- Exercise coordinates a multi-omic enrichment of sEVs in breast milk
- Exercised breast milk sEVs enhance mitochondrial respiration in UC-MSCs

## 1. INTRODUCTION

Early-life nutrition exerts a profound influence on infant growth, metabolic health, and long-term disease risk [1]. Human breast milk serves not only as a source of nutrients but is also a complex bioactive system that transmits signals to the infant which shape metabolic and immune development [2–4]. Increasing attention has focused on the capacity of breast milk to convey maternal physiological cues to the infant, raising the possibility that maternal behaviors such as exercise may influence offspring health through breast milk-borne signals [5]. Foundational work established that maternal postpartum exercise does not adversely affect breast milk macronutrient composition [6; 7], milk volume [6; 7], infant acceptance of milk [7; 8], or infant growth trajectories [7; 9], alleviating early concerns that physical activity during lactation could compromise infant nutrition. It has been demonstrated that cross-fostering mouse pups to exercise-trained dams is sufficient to improve adult offspring metabolic health, even in the absence of in utero maternal exercise exposure [10], implicating milk-borne factors as a driver of maternal exercise-induced intergenerational benefits. Together, these studies demonstrate not only that postpartum maternal exercise is compatible with successful lactation, but also that maternal exercise may promote beneficial offspring adaptations via breast milk-derived signals. However, the specific human milk components responsible for these effects, as well as the cellular mechanisms through which they act, remain largely unresolved.

Nearly two decades ago, small extracellular vesicles (sEVs; or “exosomes”) were identified as bioactive components in human milk [11]. Since then, breast milk sEVs have emerged as key candidates for mediating maternal–infant communication due to the identification of selectively packaged proteome [12], miRNA [13], and lipidome [14] cargo in breast milk sEVs. Furthermore, breast milk sEVs are resistant to digestive enzymes and acids [15], and multiple preclinical studies have demonstrated that ingested milk exosomes can survive digestion in vivo and are deposited in peripheral tissues (including the liver, GI tract, heart, lungs, brain, and others), where they exert biological effects [16–18]. These studies not only established milk exosomes as functionally relevant signaling entities (rather than inert byproducts of lactation), but have also prompted efforts to harness the potential of milk exosomes as a targeted pharmaceutical delivery system [19; 20].

Exercise is a potent systemic stimulus that, in non-lactating individuals, robustly increases circulating plasma sEVs and remodels their molecular cargo [21; 22], implicating them in inter-organ communication and metabolic adaptations [22]. Plasma sEVs are among the exercise-induced signaling molecules known as “exerkines”, which also including hormones, metabolites, and lipids which are taken up by multiple organs, and coordinate system-level metabolic adaptations [23; 24]. Thus, sEVs have been positioned as candidate mediators of the systemic benefits of exercise, capable of transmitting exercise-induced signals between tissues. Whether similar exercise-responsive signaling occurs via sEVs in breast milk, or if breast milk sEVs of exercising mothers could represent a mechanism of intergenerational communication, is not known. Breast milk sEVs are particularly well suited to mediate exercise-induced milk signaling because they can safely harbor and deliver in-tact components of translational machinery, metabolic enzymes, and regulatory miRNAs. In contrast, many of the other molecular components in breast milk, such as free metabolites or hormones, are subject to the neonatal/infant gastric pH. Thus, sEV-mediated delivery of molecular signals offers a plausible mechanism by which postpartum maternal exercise may influence infant cellular metabolism via breast milk.

In the present study, we tested the hypothesis that an acute bout of moderate-intensity aerobic exercise increases human breast milk sEV abundance and selectively remodels their molecular cargo toward pathways involved in metabolic regulation. We further hypothesized that these exercise-modified milk sEVs could exert functional effects on recipient neonatal-stage target cells.

## 2. RESULTS

### 2.1. Acute Moderate-Intensity Exercise Induces a Sustained Increase in Human Breast Milk sEVs

To determine whether acute aerobic exercise alters human breast milk sEV abundance, we employed a dietary-controlled, within-subject, sedentary time-matched design (**Figure 1A**). Healthy lactating women (**Table 1**) consumed standardized meals for two days before the initial study visit (Sedentary Day). Breast milk was collected from the second full-expression pump of the day, exactly 2 hours after a standardized breakfast. The first milk sample (Pre-Sed) was obtained prior to 90 minutes of supervised sedentary time, after which a second sample was collected (Post-Sed). The next day, participants returned (time-matched) for a baseline sample collection under the same dietary conditions (Pre-Ex) and performed 30 minutes of supervised continuous moderate-intensity (**Figure S1A**) aerobic exercise prior to milk collection at 30 minutes post-exercise (30Post-Ex; time-matched to the Post-Sed sample on the previous study visit) and 120 minutes post-exercise (120Post-Ex). sEVs were isolated by size-exclusion chromatography followed by ultracentrifugation [21], then analyzed for particle concentration, proteome, miRNAs, and acylcarnitines.

**Figure 1:**
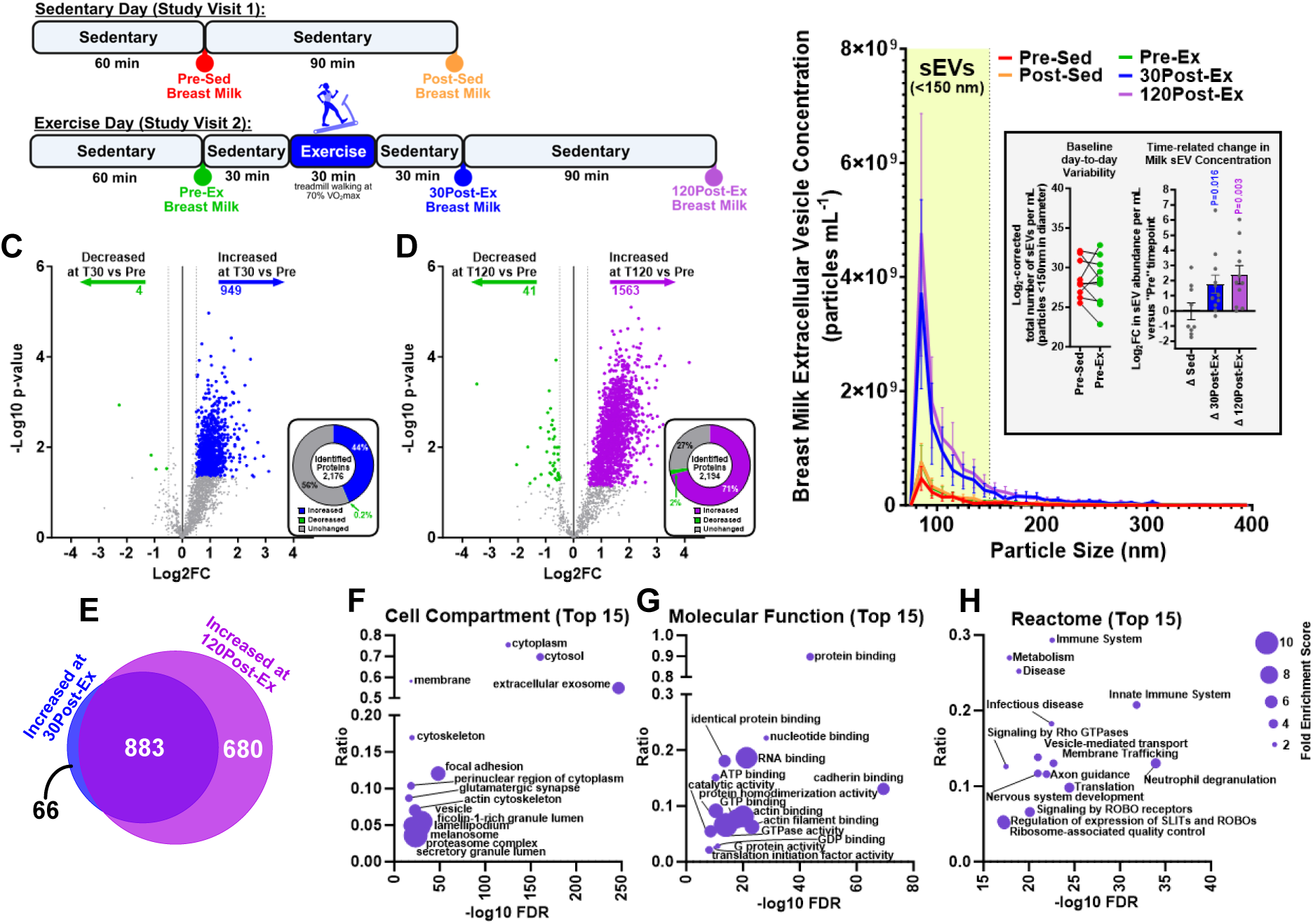
Acute Moderate-Intensity Exercise Induces a Sustained Increase in Human Breast Milk sEVs. (**A**) Study design overview. Breast milk samples were collected on sedentary (Sed) and exercise (Ex) study visits (time-matched) and small extracellular vesicles (sEVs) were isolated using size exclusion chromatography (SEC) and ultracentrifugation (UC) from fresh samples. (**B**) Nanoparticle sizing and quantification using microfluidic resistive pulse sensing from breast milk sEVs isolated by SEC (*n*=10 Pre-Sed; *n*=9 Post-Sed; *n*=11 in Pre-Ex, 30Post-Ex, and 120PostEx). Dots in the inset graphs represent individual participants. P-values were calculated using two-tailed paired t-tests. Data are presented as the mean ± SEM (**C and D**) Breast milk sEV proteomic changes at 30 (blue) and 120 (purple) minutes following a moderate-intensity exercise session versus pre-exercise (green). Unadjusted p-value is plotted on the y-axis, but only proteins with FDR-corrected (Benjamini-Hochberg) q-value of < 0.1, and |log2FC| > 0.5 are colored (*n*=11 per group). (**E**) Overlap in significantly increased (P<0.05 and FDR<0.1 and |log2FC| > 0.5) breast milk sEV proteins at 30 and 120 minutes post-exercise. (**F and H**) Bubble plots show the top 15 positively enriched Gene Ontology (Cellular Component and Molecular Function) terms and Reactome pathways in the exercise-regulated (at both 30Post-Ex and 120Post-Ex) milk sEV proteome.

**Table 1:** Breast Milk Sample Donor Characteristics. For all characteristics, *n*=11 unless specified. Because one breast milk donor gave birth pre-term, some of the pregnancy and infant characteristics are displayed without this data included (*n*=10).

| *Participant Characteristics at Study Visit (n=11)* |  |  |
| --- | --- | --- |
|  | **Age** | 31.9 ± 1.1 |
|  | **Height (cm)** | 166.8 ± 1.8 |
|  | **Weight (kg)** | 67.0 ± 1.7 |
|  | **BMI** | 24.1 ± 0.7 |
|  | **Months Postpartum at Study** | 9.4 ± 1.0 |
|  | **% Body Fat** | 35.5 ± 1.4 |
|  | **Predicted VO<sub>2</sub>max (mL kg<sup>-1</sup> min<sup>-1</sup>)** | 33.1 ± 1.6 |
|  | **Fasting Glucose (mg dL<sup>-1</sup>)** | 83.3 ± 1.9 |
|  | **Fasting Insulin (μU mL<sup>-1</sup>)** | 4.8 ± 0.8 |
|  | **HOMA-IR** | 0.99 ± 0.17 |
|  | **Race:** |  |
|  | White | 10 |
|  | Black | 1 |
| *Pregnancy and Infant Characteristics* |  |  |
|  | **Delivery Mode:** |  |
|  | Vaginal | 7 |
|  | Cesarean section | 4 |
|  | **Pre-Gravid BMI (n=10)** | 24.7 ± 0.5 |
|  | **Gestational Weight Gain (kg; n=10)** | 13.3 ± 1.3 |
|  | **Gestational Weight Gain Category (n=10):** |  |
|  | Insufficient | 3 |
|  | Appropriate | 3 |
|  | Excessive | 4 |
|  | **Gravidity** | 2.5 ± 0.4 |
|  | **Parity** | 1.5 ± 0.2 |
|  | **Infant Birth Weight (kg; n=10)** | 3.4 ± 0.2 |
|  | **Infant Sex:** |  |
|  | Male | 6 |
|  | Female | 5 |

Acute exercise robustly increased breast milk sEV particle concentration at both 30Post-Ex and 120Post-Ex versus Pre-Ex (**Figure 1B**). By contrast, particle count was not different after 90 minutes of sedentary time or between baseline samples collected on consecutive days (**Figure 1B**), indicating minimal day-to-day or time-of-day drift in milk sEV concentration. To account for variation in whole milk concentration as a potential confounder of sEV concentration, freeze dried and total milk solids were quantified across timepoints. Total solids did not differ between days, and although milk became modestly more concentrated after the first pump, this change was equivalent across exercise and sedentary conditions (**Figure S1B**). Thus, the exercise-associated increase in sEVs is not due to generalized milk concentration or maternal hydration.

Next, untargeted proteomics demonstrated that 44% and 71% of identified proteins significantly increased at 30 and 120 minutes post-exercise, respectively (**Figures 1C-D**). Because proteomics was conducted on sEVs isolated from equal milk volumes, this post-exercise proteomic enrichment likely reflects increased sEV concentration rather than changes in proteomic cargo on a per-vesicle basis. Consistent with this, normalization to total protein revealed a statistically significant proteomic shift at 120PostEx, but not at 30PostEx (**Figures S1C-D**), indicating that while sEV release is rapid and sustained, remodeling of the breast milk sEV proteome occurs over a longer post-exercise timescale. The increased sEV concentration was confirmed by significant increases in most known EV proteins at both timepoints (**Figures S1E-F**). Additionally, the sedentary control samples and day-to-day variability showed negligible proteomic shifts (**Figures S1G-H**), indicating biological specificity of the exercise-regulated sEV proteomic increase. Technical controls demonstrated that skimming milk fats and cellular debris from fresh samples prior to sEV isolation (**Figure S1I**) reduces proteomic coefficient of variation (**Figure S1J**). The greater proteomic variation in sEVs isolated from frozen whole-milk samples, potentially from apoptotic cellular debris [25], drastically impaired the detectability of exercise-induced changes in milk sEVs (**Figure S1K**), underscoring the necessity of fresh milk sample processing.

Approximately 93% of all proteins that were significantly increased at 30Post-Ex were also increased at 120Post-Ex (**Figure 1E**). Gene ontology overrepresentation analyses of these overlapping proteins showed that extracellular exosome and cytosol compartments are the most upregulated by exercise (**Figure 1F**). Furthermore, protein and RNA binding represented prominent exercise-regulated functional pathways (**Figure 1G**), indicative of greater sEV content. Reactome analyses demonstrated upregulation of immune-related pathways, consistent with an immune-enriched profile of milk sEVs [13]. These analyses support the concept that postpartum exercise induces a sustained release of breast milk sEVs.

### 2.2. Exercise-enriched sEVs in Breast Milk Contain Important Regulatory Molecular Cargo

Closer interrogation of Reactome pathway analysis for non-EV-associated processes revealed that the “translation” pathway was significantly enriched by exercise (**Figure 1H**), and 70% of exercise-regulated proteins in this pathway originated from subunits of only six major protein groups: eukaryotic translation elongation and initiation factors, large and small ribosomal proteins, and the 20S and 26S proteasome complexes (**Figures 2A and S1L**). These results suggest that postpartum exercise enrichment in milk sEVs includes key components of translational and proteo-static pathways, with potential implications for infant cellular metabolic growth.

**Figure 2:**
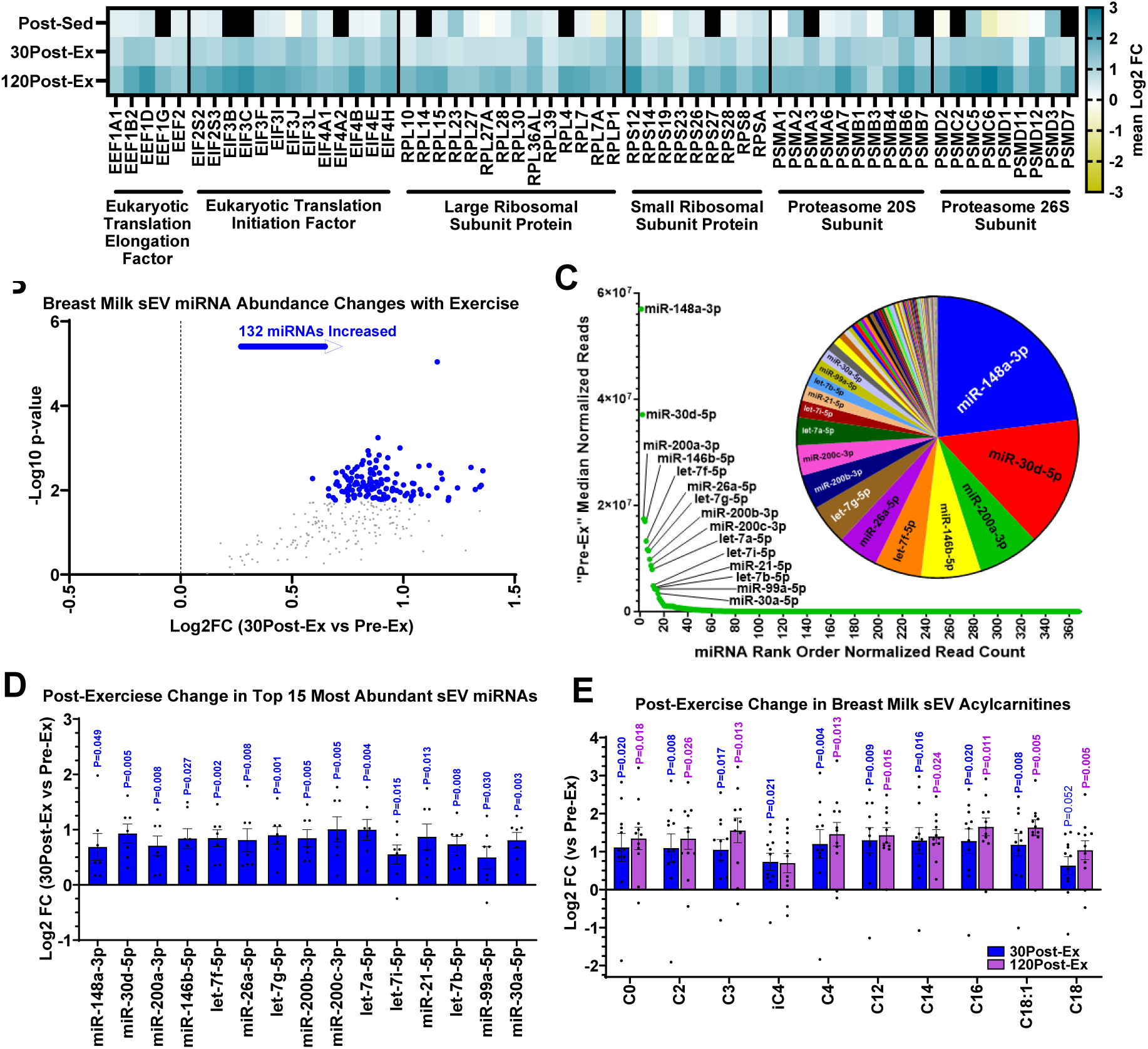
Exercise-enriched sEVs in Breast Milk Contain Important Regulatory Molecular Cargo. (**A**) Heatmap of changes in the top Reactome ‘Translation” pathway protein isoform abundances after the sedentary (Post-Sed) period (versus Pre-Sed) and 30 (30Post-Ex) or 120 (120Post-Ex) minutes following moderate intensity exercise (versus Pre-Ex). Black fill indicates a protein not detected in >50% of all samples at both timepoints. (*n*=9 or Post-Sed comparisons, *n*=11 or Post-Ex comparisons). (**B**) Change in the normalized abundance of breast milk sEV miRNAs following a moderate-intensity exercise session versus pre-exercise (*n*=7). Unadjusted p-value is plotted on the y-axis, but only miRNAs with P<0.05, FDR-corrected (Benjamini-Hochberg) q-value of < 0.1, and |log2FC| > 0.5 are colored. (**C**) Baseline (Pre-Ex) median miRNA abundances (based on normalized reads) are plotted in rank order and the top 15 miRNAs are labeled in the waterfall plot (*n*=7). Pie chart represents that the top 15 miRNAs (labeled) account for ∼86% of the total breast milk sEV miRNA content. (**D**) Postexercise changes in the top 15 most abundant breast milk sEV miRNAs. Dots represent individual participants. Unadjusted P-values were calculated using two-tailed paired t-tests between 30Post-Ex versus Pre-Ex values (*n*=7). (**E**) Post-exercise changes in breast milk sEV acylcarnitine species. One-way ANOVA was used to test the time-related change in breast milk sEV acylcarnitines on the exercise visit, and p values represent significant changes at 30Post-Ex (blue) and 120Post-Ex (purple) versus Pre-Ex. (*n*=11). The quantified data in (D)-(E) are presented as the mean ± SEM, and dots represent individual participants.

Breast milk sEVs also contain miRNAs with regulatory functions for recipient cells [26]. We found that total RNA (including small RNAs) abundance increased at 30Post-Ex across all 11 participants (**Figure S2A**), paralleling the rise in sEV concentration. In addition, miRNA sequencing revealed a significant (FDR-corrected) post-exercise increase in 132 of the 305 reliably quantified breast milk sEV miRNAs (**Figure 2B**) when expressed relative to RNA isolated per volume of milk. Since the absolute miRNA material in sEVs is relatively low, higher abundant miRNAs are the most likely to exert a meaningful effect on target cells. Sorting miRNAs based on normalized read count revealed that only 15 miRNAs accounted for 86% of the breast milk sEV miRNA content (**Figure 2C**). Most of these highly abundant miRNAs approached or surpassed an exercise-induced Log2 fold change of 1, representing near-doubling of deliverable miRNA content in breast milk sEVs (**Figure 2D**). Notably, many of the most abundant human milk miRNAs from the current study (including miR-148a-3p, miR-30d-5p, miR200a-3p, miR-146b-5p, let-7f-5p, let7a-5p, miR30a-5p, and miR-21-5p) are known to promote metabolism, immune function, neurogenesis, and intestinal maturation [27].

Human breast milk sEVs also contain metabolites important for cellular metabolism and molecular signaling [28]. A previous report demonstrated that whole milk long-chain acylcarnitines, and other metabolites, are altered by acute exercise [29]. In the current study, 10 acylcarnitine species in breast milk sEV preparations were reliably quantified, nearly all of which significantly increased at 30Post-Ex and 120Post-Ex (**Figure 2E**). Notably, no time-dependent changes were observed in sEVs for any acylcarnitine species during the sedentary control visit (**Figures S2B-C**), and baseline concentrations did not differ between study days (**Figure S2D**), reaffirming that increases in breast milk sEV acylcarnitines are attributable to the acute exercise intervention. Although exercise-induced increases in multiple long-chain acylcarnitines in whole milk were observed (**Figure S2E**), similar increases on the sedentary control visit were also detected (**Figure S2F**). Thus, the magnitude of changes in whole milk metabolite concentrations in the hours post-exercise is confounded by a diurnal effect on whole milk composition (**Figure S2G**), despite minimal day-to-day variability in whole milk acylcarnitines (**Figure S2H**). These results suggest that the sEV compartment represents a biologically informative and exercise-responsive fraction of milk, in contrast to whole milk, where diurnal changes may influence acute exercise effects.

### 2.3. Breast Milk sEVs from Exercised Mothers Enhance Cellular Metabolic Capacity

To determine whether breast milk sEVs can transmit “exerkine-like” signals into functional effects on recipient cells, we exposed milk-derived sEVs obtained before and after maternal exercise to primary human umbilical cord–derived mesenchymal stem cells (UC-MSCs). UC-MSCs are a physiologically relevant, developmentally immature cell model capable of responding to extracellular metabolic signals [30]. Following a 48-hour incubation with sEV particles isolated from an equal volume of skimmed milk at Pre-Ex, 30Post-Ex, or 120Post-Ex, UC-MSC mitochondrial capacity and content were assessed (**Figure 3A**) at approximately 100% confluence (**Figure 3B**). A similar rate of growth was observed in UC-MSCs exposed to sEVs isolated either before or after exercise (**Figure S3A**). However, UC-MSCs exposed to post-exercise breast milk sEVs displayed a greater maximal mitochondrial respiratory capacity compared to UC-MSCs exposed to breast milk sEVs isolated before exercise (**Figures 3C and S3B**). The enhanced oxidative capacity of UC-MSCs exposed to breast milk sEVs isolated post-exercise did not appear to be due to greater mitochondrial content, based on unaltered Complex II, III, IV, and V abundances (**Figures 3D-E**) or citrate synthase activity (a canonical marker of mitochondrial content) (**Figure 3F**). Together, these findings suggest that greater post-exercise concentration of breast milk sEVs enhances intrinsic mitochondrial respiratory capacity in recipient cells without increasing mitochondrial content.

**Figure 3:**
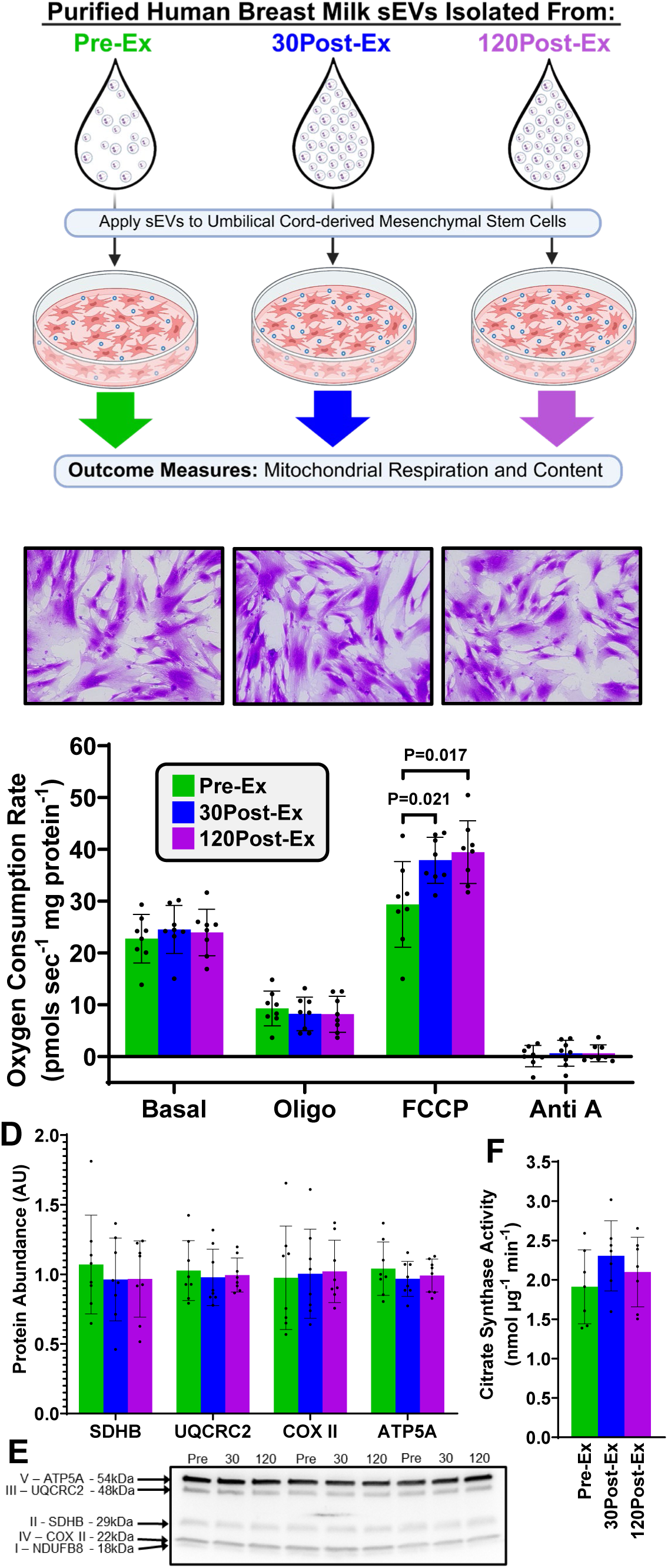
Breast Milk sEVs from Exercised Mothers Enhance Cellular Metabolic Capacity. (**A**) Experimental Design. sEVs isolated by size exclusion chromatography from skimmed breast milk obtained before (Pre-Ex), 30 minutes after (30Post-Ex), and 120 minutes after (120Post-Ex) a bout of postpartum exercise were pelleted and resuspended in cell culture media, then applied to human umbilical cord-derived mesenchymal stem cells (UC-MSCs). (**B**) Representative images of crystal violet stained UC-MSCs after 48 hours of sEV treatment (x20 magnification). (**C**) Oxygen consumption rate of UC-MSCs exposed to breast milk sEVs isolated at Pre-Ex, 30Post-Ex, and 120Post-Ex in the basal state and in the presence of oligomycin (2 µg/ml; Oligo), FCCP (0.25µM; Carbonyl cyanide p-trifluoro-methoxyphenyl hydrazone), and Antimycin A (2.5 µM; Anti A). (**D and E**) Quantified abundance of mitochondrial complex proteins from UC-MSCs exposed to breast milk sEVs isolated Pre-Ex, 30Post-Ex, and 120Post-Ex, and representative immunoblots. (**F**) Citrate synthase activity in UC-MSCs exposed to breast milk sEVs. For the quantified data in (C), (D), and (F), Dots represent data from an individual sample. Statistical significance was calculated using one-way ANOVA followed by multiple comparisons (*n*=8 per group). P-values are presented on the graph. Data are presented as the mean ± SEM.

## 3. DISCUSSION

The current study demonstrates that postpartum exercise increases breast milk sEV abundance and drives multi-omic remodeling of sEV cargo with functional cellular consequences. The most important findings were that: 1) a single bout of moderate-intensity exercise increases breast milk sEV concentration for multiple subsequent milk expressions, 2) postpartum exercise coordinated a multi-omic enrichment of sEVs in breast milk, and 3) breast milk sEVs obtained post-exercise enhance mitochondrial respiration in neonatal-stage target cells compared with pre-exercise sEVs. Together, these results identify postpartum maternal physical activity as a practical intervention capable of modulating maternal–infant metabolic signaling via breast milk sEVs.

The convergent multi-omic changes support a model in which exercise increases the biological payload of sEVs per volume of breast milk. Notably, this is evident by 30-minutes post-exercise. In contrast to plasma, where sEV half-life is ∼2 minutes [31], breast milk sEVs remained substantially elevated even at the second post-exercise milk collection (120Post-Ex). Because plasma sEV concentrations normalize within ∼30-90 minutes post-exercise [21; 32; 33], we anticipated that milk sEVs from the second post-exercise pump would no longer reflect the exercise stimulus. Instead, our findings suggest compartmental differences between plasma and breast milk sEVs. Although we cannot determine the cellular origin of exercise-responsive milk sEVs in humans, their sustained elevation, combined with recent murine data that >80% of milk sEVs are mammary epithelial cell-derived [34], points to a predominantly mammary origin. Thus, our data suggest that mammary epithelial cells retain a transient “memory” of acute exercise, which sustains sEV production well after the cessation of exercise. This apparent “memory element” of the exercise stimulus is reminiscent of a well-described phenomena in skeletal muscle, where a single bout of exercise enhances insulin-stimulated glucose uptake for multiple days [35]. Although the molecular basis of sustained exercise responsiveness remains incompletely understood in both muscle and mammary cells, prolonged sEV production following exercise in the lactating mammary gland has important functional implications.

The central translational message of our findings is feasibility. The intervention in the current study was moderate intensity walking, rather than vigorous exercise, making the behavior broadly achievable for postpartum women. Furthermore, because the sEV increase persists across multiple milk expressions, a single walking session may enhance the signaling environment experienced by an infant across several feeds, an important consideration for mothers unable to exclusively breastfeed. In practice, this could mean that partial breastfeeding coupled with maternal walking confers measurable biological advantages to the infant, providing infant-centered maternal motivational value. Among many of the important next questions is whether exercise intensity (light vs moderate vs vigorous), modality (aerobic vs resistance), and/or duration produce qualitatively different responses. Furthermore, this study cohort consisted of healthy women, as reflected by BMI, percent body fat, HOMA-IR, and aerobic capacity. Whether similar responses occur in mothers with obesity or diabetes, as their infants are at greater risk for metabolic disease [36], remains an important question for future investigation. Our observation that exercise enriches milk with sEVs containing translational and metabolic cargo capable of altering recipient cell function therefore highlights a potential mechanism by which maternal exercise could shape early-life metabolic programming in at-risk neonates and infants.

Previous studies consistently demonstrate that maternal exercise before and/or during pregnancy improves rodent offspring glucose tolerance [37–39], and that maternal-fetal exercise-mediated communication occurs directly through placental-secreted factors [40]. Human studies demonstrate that maternal physical activity during pregnancy is associated with lower risk of macrosomia and childhood obesity [41; 42], potentially due to greater infant metabolic rate [43; 44]. Moreover, gestational exercise is linked to enhanced mitochondrial respiration in UC-MSCs [30]. Thus, a strong precedent exists for maternal exercise-associated signaling during gestation to influence offspring health, though most previous studies have focused on gestational exercise rather than postpartum, preventing the ability to discern is maternal exercise transmits benefits to offspring via breast milk. Evidence for milk-borne factors as a contributor of maternal exercise-induced intergenerational metabolic benefits have been demonstrated in mice either by cross-fostering offspring to exercise trained dams exposure [10] or exercise training dams in the post-partum period [45]. However, the molecular response of human breast milk to postpartum exercise, and the cellular mechanisms through which these signals exert metabolic effects, remain largely unresolved. The current study provides the first proof-of-principle evidence in humans that postpartum maternal exercise provides metabolically beneficial signals via increased breast milk sEV concentration. We captured the endogenous regulatory capacity of acute exercise on human milk sEV concentration, then tested if this physiological increase in sEVs resulted in meaningful metabolic effects when applied to perinatal human cells. Rather than relying on ex vivo concentration or supraphysiologic dosing paradigms, we revealed that a biologically relevant increase in milk sEV concentration can impact cellular function. Identifying the specific milk sEV signaling mechanisms driving postpartum maternal exercise effects could inform the development of formula supplements designed to recapitulate beneficial sEV functions. Within the biopharmaceutical industry, there are currently multiple companies aimed at harnessing the potential of milk sEVs as a targeted delivery systems [19; 20] [46]. Our findings suggest that maternal physical activity may serve as a behavioral analogue to these approaches, harnessing endogenous biology to enhance milk-derived signaling for the infant.

Here, we studied the sEV fraction of breast milk because other human studies to date have provided no clear exercise-responsive candidate molecules in whole milk. Postpartum exercise in humans alters breast milk adiponectin and insulin concentrations [47; 48], but these peptide/protein hormones will likely degrade in the low neonatal/infant stomach pH, which despite being relatively neutral at birth [49] drops to < 3 by 2-weeks of age [50], preventing peptide hormones from reaching the infant circulation intact. In contrast, breast milk sEVs are resistant to digestive enzymes and acids [15], are deposited in peripheral tissues, and exert biological effects [16–18]. Although several milk metabolites, such as the oligosaccharides 3′-sialyllactose (3’SL) and 6′-sialyllactose (6’SL), are relatively resistant to digestion [51], they are not significantly altered in human breast milk by acute aerobic exercise compared to time-matched sedentary controls [52]. Recent work demonstrated that 28 metabolites [29] and the lipokine 12,13-diHOME [53] were significantly altered by moderate-intensity aerobic exercise, but the absence of time-matched sedentary control samples in these studies limits interpretability. Thus, although non-sEV compartments of whole milk are an important source of nutrients, the current study is groundbreaking because milk sEVs are currently the only acute exercise-regulated signaling entity in breast milk with a demonstrated functional link for recipient neonatal/infant cells.

This study identifies breast milk sEVs as a previously unrecognized exercise-responsive signaling system during human lactation. A single bout of moderate-intensity postpartum maternal walking not only increases sEV abundance but also enriches their molecular cargo and functional impact on neonatal-stage cells. Together, these findings reveal a previously unrecognized pathway through which maternal postpartum behavior may shape early-life metabolic programming.

## 4. MATERIALS AND METHODS

### 4.1. Experimental Model and Study Participant Details

#### 4.1.1. Study Approvals and Overall Design

The study design was approved by the Mayo Clinic Institutional Review Board and registered under Clinical Trials (ClinicalTrials.gov Identifier NCT06892483). All participants were informed of the study procedures and provided written consent. Participants underwent one screening visit and two outpatient study visits that included a dual-energy X-ray absorptiometry (DEXA) scan, an aerobic capacity test, a subsequent moderate-intensity exercise session, blood draws, and breast milk collections. A total of 11 lactating women completed all study procedures.

#### 4.1.2. Screening Visit

Participants were recruited using word of mouth and posted flyers in lactation rooms across the Mayo Clinic campus in Rochester, MN. Potential participants contacted the study team and completed a phone-screening questionnaire, then eligible women were scheduled for an in-person screening visit. The screening visit included meeting with a study team member who described all procedures, then participants provided written consent. A fasting blood sample was collected to assess plasma glucose, insulin, and comprehensive chemistry panel (including CBC, serum creatinine, liver function tests and TSH, and lipid panel). A general medical examination was performed (including vital signs) and a urine sample was obtained to verify that participants were not pregnant at the time of testing. Then, participants met with a registered dietician to discuss food preferences for weighed-meals followed by body composition assessment using dual-energy X-ray absorptiometry (DEXA) and a submaximal VO_2_ test on a treadmill using a modified Balke protocol to estimate VO_2_max. Submaximal VO_2_ testing included continuous measures of indirect calorimetry (Parvo) and heart rate with 12-lead electrocardiogram (Quintin). Manual blood pressures were also collected. The incremental exercise test began at 0% grade and a participant-selected walking pace between 3-3.5 mph. Treadmill speed was kept constant throughout the test and grade was increased by 2.5% at the end of each 2-minute stage. The test was terminated when participants reached 90% of their age-predicted max heart rate. Predicted VO_2_max, calculated using the linear relationship between heart rate and VO_2_, was used to extrapolate the measured VO_2_ to the age-predicted max heart rate.

#### 4.1.3. Inclusion/Exclusion Criteria

Inclusion criteria were maternal age of ≥ 18 years, ≥ 2 months postpartum, actively lactating and capable of producing > 3 oz of breast milk per pumping session, singleton pregnancy. Exclusion criteria included maternal BMI ≥ 30 kg m^−2^, fasting glucose ≥ 100 mg dL^−1^, cardiovascular disease, abnormal renal function (GFR < 60 mL min^−1^ 1.73 m^−^²), abnormal liver function (AST > 2 time the normal upper limit), uncontrolled thyroid disease (TSH undetectable or >10 mIU L^−1^), uncontrolled arterial hypertension (resting diastolic blood pressure >90 mmHg and/or systolic blood pressure >160 mmHg), positive pregnancy test, active smoking or tobacco use, or alcohol abuse.

#### 4.1.4. Outpatient Study Visits

Within 60 days following the initial screening visit, eligible participants returned to the Clinical Research Unit (CRU) at Mayo Clinic for two outpatient study visits on back-to-back days. Prior to the first outpatient visit, participants consumed 2-days of weighed meals (20% protein, 50% carbohydrates and 30% fat) to minimize fluctuations in body weight. Caloric requirement was estimated using the Müller equation [54], which is the most accurate equation for predicting resting energy expenditure (REE) in lactating women [55], ensuring standardized, accurate, and abundant nutrients prior to the study visit. After waking in the morning, participants breastfed their child, then fully pumped both breasts, and immediately ate a standardized breakfast (25% of daily kcals) while at home. Participants arrived at the CRU within 1 hour of ingesting their breakfast for the first outpatient study visit. Upon arrival, participants remained sedentary for an additional 1 hour (or more if they arrived early) prior to fully pumping both breasts. This milk collection was exactly 2 hours after ingesting breakfast. Milk from both breasts was combined and gently mixed. Approximately 50 mL of the combined breast milk was aliquoted for subsequent experimentation. The remainder was returned to the participant. A fresh 20 mL aliquot was used for isolation of milk sEVs (see below) immediately after collection, and the remaining 30 mL of whole milk was aliquoted and frozen in 2mL low-bind tubes. This milk collection timepoint was termed “Pre-Sed”, because after milk collection the participants remained sedentary in the CRU for an additional 90 minutes with ad libitum access to water. No food or liquid calories were consumed at this time. Participants then fully pumped both breasts and milk was collected as stated above. This timepoint was termed “Post-Sed”. Participants were then provided with standardized meals for the remainder of the day and the following morning’s breakfast. On the following morning, participants repeated the same morning procedure (feed/pump and breakfast) prior to arriving at the CRU for the second outpatient visit. Participants again remained sedentary for at least 60 minutes, and milk collection was performed at 2 hours post-breakfast. This timepoint was termed “Pre-Ex”, because 30 minutes after milk collection, the participants performed 30 minutes of inclined treadmill walking at approximately 70% of predicted VO_2_max based on submaximal VO_2_ testing on the screening day. Rating of perceived exertion (RPE; Borg) was recorded every 10 minutes throughout the test to confirm the participants exercised at a moderate intensity, according to ACSM guidelines. At the end of the aerobic exercise session, participants were again asked to remain sedentary for 30 minutes, then fully pumped both breasts for milk collection. This timepoint was termed “30Post-Ex”. The participants remained sedentary for an additional 90 minutes and then one final milk collection was performed at 120 minutes-post-exercise (“120Post-Ex”).

### 4.2. Isolation of Small Extracellular Vesicles

20mL of fresh whole milk was centrifuged in low bind tubes at 3,000 x g for 10 minutes at room temperature. The top fatty layer was perforated with a sterilized spatula to allow for removal of the skimmed milk layer with a pipet without mixing the fatty layer and skimmed milk supernatant layer. The skimmed milk fraction was transferred to new low-bind tubes and centrifuged at 10,000 x g for 10 minutes at room temperature. The skimmed milk supernatant fraction was again removed and transferred to a new low-bind tube. If a thin fatty layer was visible after centrifugation, then an additional 10-minute spin at 10,000 *g* was performed followed by supernatant removal until fatty layer was no longer visible. After centrifugation steps, approximately 10 mL (depending on lipid proportion of sample) of skimmed milk supernatant remained. Freshly skimmed milk supernatant was applied to 70 nm size-exclusion chromatography (SEC) columns (Izon Science, Christchurch, New Zealand) and the flow-through, containing sEVs in 1X PBS, was collected in low bind tubes, and portions were aliquoted for various down-stream assays including particle counting and cell culture studies (see below). The remainder was aliquoted in tubes and immediately ultracentrifuged (UC) for 90 minutes at 140,000 *g* at 4°C.

### 4.3. Quantification of Particle Size and Concentration

nCS1 (Spectradyne, Signal Hill, CA) was used to conduct microfluidic resistive pulse sensing (MRPS) measurements of particle size distribution (PSD) from the SEC eluate of skimmed breast milk. A solution of 1% Tween 20 (v/v) in PBS was prepared to prime the instrument and to dilute each sample prior to analysis. Samples were initially diluted (1:2-1:2000) in PBS + 0.1% Poloxamer 188 prior to analysis. The instrument parameters (e.g. pressure at each port of cartridges) were determined as previously described [56; 57]. 3 μl of each sample was loaded onto C-400 (65 to 400 nm) polydimethylsiloxane cartridges to determine the PSD of particles in each sample. For post-acquisition analysis nCS1 Data Viewer (Version 3.5.0.67, Spectradyne) was used. The peak filters were applied to all single particles individually to exclude false positive signals. All raw data were coalesced after processing by applying additional filters and subtracting background noise, as recommended by the manufacturer, and suggested previously [56]. After manual data analysis, PSDs were exported to excel files with 10-nm bins.

### 4.4. Sample Preparation for Proteomic Analysis of Small Extracellular Vesicles

sEV pellets used for proteomics analysis were isolated from 500µl of fresh skimmed milk by SEC-UC (described above). sEV pellets were denatured directly in UC tubes using 45 µL sodium dodecyl sulfate (SDS) lysis buffer (5% SDS, 50 mM TEAB at pH 7.6, in LC-MS-grade water), vortexed, and then transferred to a low-bind tube. An additional 15 µL of lysis buffer was added to the empty UC tube to collect any remaining sample, vortexed, and then transferred to the low-bind tube for a total volume of 60 µL. Next, 2µL of reductant (120 mM TCEP, in LC-MS-grade water) was added, samples were briefly vortexed, and incubated on a heat block at 55°C for 10 minutes. Samples then were cooled at room-temperature for 20 minutes before adding 2 µL of alkylator (500 mM IAA, in LC-MS-grade water). Samples were vortexed and left at room temperature for 10 minutes. Next, 5 µL of acidifier (27.5% phosphoric acid, in LC-MS-grade water) was added, and then 350 µL of binding/wash solution (100 mM TEAB at pH 7.6, in 90% LC-MS-grade methanol) was added. Samples were inverted twice (not vortexed), and the entire sample was loaded into a well of a S-Trap™ 96-well Mini Plate (P002-96MNT, Protifi, Fairport, NY, USA). The plate (with all samples loaded) was centrifuged at 2,000 *g* for 2 minutes at room temperature. Flow through was discarded and washing was repeated 3 times by adding 250 µL of binding/wash solution to each well followed by centrifugation at 2,000 x g for 2 minutes. Following the final wash, the S-Trap plate was moved atop a new 96-well low-bind collection plate, and 125 µL of digestion solution (50 mM TEAB at pH 8.0 in LC-MS-grade water) containing 0.55 µg of trypsin/Lys-C per 125 µL was added to each well. The sample plate was incubated (loosely covered to minimize evaporative loss) in a humidified chamber at 47 °C for 1 hour. An additional 80µL of digestion solution (without trypsin/Lys-C) was added, followed by an additional 1 hour of humidified incubation at 47 °C. Samples were eluted by three sequential additions of 80 µL of digestion solution (without trypsin-Lys-C), elution solution 1 (0.2% LC-MS-grade formic acid in LC-MS-grade water), and elution solution 2 (50% LC-MS-grade acetonitrile in LC-MS-grade water) followed by centrifugation (2,000 *g* for 2 minutes) after each 80 µL addition. Lastly, 400 µL of the final sample volume was transferred to glass autosampler vials, frozen at -80°C, and dried using SpeedVac with centrifugation.

### 4.5. Mass Spectrometry

Prepared peptide samples were subject to mass spectrometer data acquisition using a Thermo Ultimate 3000 RSLC nano HPLC autosampler system coupled to an Orbitrap Fusion Lumos mass spectrometer (Thermo Fisher Scientific, Waltham, MA). Each experiment included multiple biological replicates, corresponding to individual participants, with one technical replicate per sample (n = 1). Peptides were loaded onto a 330 nL Halo 2.7 μm ES-C18 EXP stem trap (Optimize Technologies, Oregon City, OR). Chromatography was performed using a 2%–30% gradient of solvent B over 60 min and up to 40% B over the next 10 minutes, where solvent A is (98% water/2% acetonitrile/0.2 % formic acid) and solvent B is (80% acetonitrile/10% isopropanol/10% water/0.2% formic acid). Peptides were eluted at a flow rate of 250 nL min^−1^ from the trap through a Pico Tip emitter 75 µm x 30 cm column (New Objective, Littleton, MA) hand packed with Acclaim Pepmap C18 resin (Thermo Fisher Scientific, Waltham, MA). Orbitrap Fusion Lumos was set to acquire ms1 survey scans from 340–1600 m/z at resolution 120,000 (at 200 m/z) with an AGC target of 4e5 ions. Survey scans were followed by HCD (30% collision energy) ms2 scans in the ion trap with an AGC target of 1e4 ions and maximum ion injection time of 35 msec and a mass range of 200-1200 m/z. The isolation window was set at 1.5 Da, both exclude isotopes and scan only single charge states were set to true, and dynamic exclusion placed selected ions on an exclusion list for 60 s.

### 4.6. Mass Spectrometry Analysis

Mass spectrometry generated data files were first analyzed by Proteome Discoverer software (v2.5, Thermo Scientific) against a human SwissProt FASTA database (release 2025-04-23) appended with common contaminants using the Sequest HT search engine [58]. The precursor mass tolerance was set to 10 ppm and fragment tolerance at 0.02 Da. Using a decoy database, false discovery rate of 1% was set for both peptides and their resulting proteins. Enzyme specificity was set to trypsin with a maximum of two missed cleavages allowed. Protein and peptide data was exported from the Proteome Discoverer result files for additional relative quantitation calculations. Briefly, peptide and protein intensities were log2 transformed and median normalization was applied to the experimental samples. An ANOVA test was used to detect differentially expressed proteins and peptides between groups of interest, of which p-values were FDR corrected using the Benjamini-Hochberg procedure. For the final analysis, only proteins that were detectable across all timepoints (for any given statistical comparison) in at least 6 participants were used. All raw data will be publically available upon manuscript acceptance in a peer-reviewed journal.

### 4.7. Proteome Pathway Analysis

Functional enrichment analysis of the breast milk sEV proteome was performed using DAVID Bioinformatics Resources (Database for Annotation, Visualization and Integrated Discovery) [59]. Proteins identified as significantly increased at both 30PostEx and 120PostEx were mapped to their corresponding gene symbols and submitted as an unranked gene list. Overrepresentation analysis was conducted using default parameters against curated annotation categories, including Gene Ontology (GO) Molecular Function, GO Cellular Component, and Reactome Pathway Database. Enrichment significance was determined using a modified Fisher’s exact test (EASE score) implemented within DAVID. Because the analysis was performed using an unranked gene list, no weighting by fold-change magnitude was applied, and results therefore reflect the presence of enriched functional themes rather than quantitative differences in pathway activity. Multiple testing correction was applied using the Benjamini–Hochberg method, and pathways were considered significantly enriched at a false discovery rate (FDR) threshold of <0.05.

### 4.8. Analysis of Acylcarnitines in Small Extracellular Vesicles and Whole Milk

Acylcarnitines (specifically C0–C18:1) were measured by LC-MS as previously described [60; 61], with slight modifications. Briefly, 1 mL of breast milk supernatant was isolated by SEC (described above) and pelleted by ultracentrifugation to isolate sEVs. The breast milk sEV pellet was spiked with an internal standard consisting of isotopically labeled acylcarnitines. For whole milk analysis, 25 µl of thawed whole milk was spiked with internal standard. The samples were then extracted with cold MeOH:DCM (1:1) followed by centrifugation at 18,000 *g* for 15 minutes. The supernatant was transferred to another vial, dried down and reconstituted in running buffer. A calibration curve was made from an acylcarnitine mix aliquoted at various concentrations and spiked with the same internal standard as the samples. The samples and calibration standards were analyzed on a Thermo Quantiva triple quadrupole mass spectrometer coupled with a Waters Acquity liquid chromatography system. Data acquisition was done using selective ion monitoring (SRM). The analyte concentrations of each unknown were calculated against their perspective standard curves. Values for multiple sEV samples fell below the level of detection for iC5, C5, C8, C10, C4-OH, and iC5-OH, and therefore these data are not presented. All other acylcarnitine species measured in sEVs fell above the level of detection and were quantifiable. Values from multiple whole milk samples for C16 and C18 fell below the level of quantitation and therefore data from these acylcarnitine species are not shown.

### 4.9. Analysis of miRNAs in Small Extracellular Vesicles and Whole Milk

sEV pellets used for miRNA analysis were isolated from 3mL of fresh skimmed milk by SEC-UC (described above). Total and small RNAs were isolated from sEV pellets using QIAGEN’s miRNeasy Micro Kit. Disruption of sEV membranes and homogenization of RNA was achieved using QIAzol and vortexing. The supplied phenol-based protocol without on-column DNase digestion or miRNA purification was used to elute RNA in 14ul of RNase-free water. Isolated total and small RNA from breast milk sEVs were sent to Azenta Life Sciences (South Plainfield, NJ, USA) for primary quality control analysis and miRNA next-generation sequencing. Total and small RNA concentrations for quality control were measured using a Qubit analyzer (Thermo Fisher Scientific, Waltham, MA) and revealed that samples from 4 of the 11 participants did not have adequate RNA content for reliable analysis. We present the total and small RNA concentration data in sEVs from all 11 participants, but miRNA-sequencing analysis was only reliably conducted on samples from 7 participants. Small RNA libraries were prepared using adapter ligation chemistry selective for RNAs bearing 5′ phosphate and 3′ hydroxyl termini (consistent with Dicer-processed products), followed by sequencing using an Illumina platform with paired-end 2×150 bp reads. Raw reads were adapter-trimmed and quality-filtered, followed by alignment to miRBase for known miRNA identification. Remaining reads were mapped to the reference genome for novel miRNA prediction based on secondary structure analysis. Since miRNA read counts are reflective of an equal amount of total and small RNA loaded for sequencing analysis, miRNA read counts were normalized to the total and small RNA concentrations measured at quality control analysis. Thus, this normalization enables timepoint comparison (Pre-Ex vs 30Post-Ex) of sEV miRNA content relative to milk volume, which is more physiologically relevant in the current study than comparing sEV miRNA content relative to sEV total and small RNA concentration. Only miRNAs that were present in both Pre-Ex and 30Post-Ex samples from at least 6 of the 7 participants are presented.

### 4.10. Whole Milk Total Solids Concentration

To measure whole milk solids concentration, 1.8 ml of frozen whole milk was lyophilized in pre-weighed glass tubes. After Lyophilization, tubes with dried milk were weighed and dried sample weight (in mg) was made relative per volume (in mL) of whole milk.

### 4.11. Cell experiments

Primary human umbilical cord-derived mesenchymal stem cells (UC-MSCs) from a single donor were obtained in passage 2 from ATCC (Manassas, VA, USA; Cat# PCS-500-010). Cells were seeded onto 6-well plates (Corning, Corning, NY, USA) at a density of approximately 3 × 10^5^ cells cm^−2^ in mesenchymal growth media: low-glucose DMEM (Gibco Laboratories, Grand Island, NY, USA; Cat# 11885084), 10% MSC-qualified fetal bovine serum (Gibco Laboratories, Grand Island, NY, USA; Cat# 12661-002), and a 1× mixture of gentamicin (10ug/ml) amphotericin B (0.25ug/ml) (Life Technologies, Gaithersburg, MD, USA; Cat# R01510). All experiments were conducted on UC-MSCs in passage 4 after one cryopreservation/rescue between passage 2 and 3.

#### 4.11.1. Suspension of sEVs in Cell Culture Media

Breast milk sEVs were isolated from 1 mL of freshly skimmed milk by SEC, yielding 3.2 mL of purified sEVs in 1X PBS, which were stored at -80 °C. On the day of treatment, sEV samples were thawed and transferred to ultracentrifuge tubes under sterile conditions. Samples were centrifuged at 140,000 *g* for 90 minutes at 4 °C. Following centrifugation, PBS was carefully removed, and sEV pellets were resuspended in 4.4 mL of cell culture media, which included vortexing for 20–30 seconds. Freshly resuspended sEVs were then applied to UC-MSCs at approximately 80% confluence for 48 hours. Accounting for SEC dilution and subsequent ultracentrifugation and resuspension, each 1 mL of final sEV-containing media represented vesicles derived from approximately 165 µL of the original skimmed milk. In the Pre-Ex (sedentary) samples, microfluidic resistive pulse sensing quantified ∼1.7 × 10^9^ particles per mL of eluate (Fig. 1b). Multiplying this value by 3.2/4.4 (representing the eluate to media dilution), this corresponds to ∼1.2 × 10^9^ particles per mL of Pre-Ex treatment media. Since sEVs are more concentrated in milk postexercise, we calculated 30Post-Ex and 120Post-Ex treatment medias to contain ∼5.6 × 10^9^ and ∼7.3 × 10^9^ particles per mL of treatment media, respectively. Cells were treated with 1 mL of sEV-containing media per well in a 6-well plate. Assuming >1.0 × 10^6^ cells per well at confluence, this corresponds to an estimated exposure of approximately 1200, 5600, and 7300 sEV particles per cell for Pre-Ex, 30Post-Ex, and 120Post-Ex treatment conditions, respectively. This range of sEV concentrations is consistent with doses reported to elicit source-specific biological effects, rather than nonspecific immune responses associated with supraphysiologic exposure [62].

#### 4.11.2. Crystal Violet Staining

To enable visualization of UC-MSC cell density and viability, crystal violet staining was conducted on samples exposed to sEVs isolated Pre-Ex, 30Post-Ex, and 120Post-Ex. Following treatment, cells were washed gently with 1X PBS and fixed in 10% formalin PBS for 15 minutes on an orbital shaker at room temperature. Wells were then stained with 0.05% (w/v) crystal violet solution for 15 minutes. Excess stain was thoroughly washed with ddH_2_O, and plates were air-dried. For qualitative visual assessment, stained cells were imaged using brightfield microscopy. For quantitative analysis, bound dye was solubilized using 10% acetic acid on an orbital shaker for 30 minutes, and absorbance was measured at 590 nm.

#### 4.11.3. O2K Respirometry

High-resolution O_2_ consumption measurements were conducted using the Oroboros Oxygraph-2K (Oroboros Instruments, Innsbruck, Austria) in intact cells in mesenchymal growth media. After recording the basal respiration, ATP-synthase (Complex V) was inhibited by the addition of oligomycin (2 mg ml^−1^). Maximal respiratory capacity was then assessed by addition of sequential titrations (0.25–1.0 μM) of the uncoupler FCCP (Carbonyl cyanide p-trifluoro-methoxyphenyl hydrazone, C10H5F3N4O). Finally, Antimycin A (2.5 μM) was added to block the electron transport chain and measure non-mitochondrial oxygen consumption. Protein content from UC-MSCs used for the respiration assay was determined using the DC Protein Assay (Bio-Rad Laboratories, Hercules, CA, USA).

#### 4.11.4. Citrate Synthase Activity

Citrate synthase activity was measured in 96-well plates on a SpectraMax Plus 384 (Molecular Devices LLC, Sunnyvale, CA) and is often associated as an index of mitochondrial content. Briefly, following protein quantification as previously described [63], equal amounts of protein from cell lysates were used to determine enzyme activity spectrophotometrically by monitoring the formation of 5-thio-2-nitrobenzoic acid (TNB) at 412 nm following the reaction of acetyl-CoA with oxaloacetate in the presence of 5,5′-dithiobis-(2-nitrobenzoic acid) (DTNB) in potassium phosphate buffer. Citrate synthase activity was calculated from the linear rate of change in absorbance, normalized to total protein content, and expressed as nmol μg protein^−1^ min^−1^.

#### 4.11.5. Immunoblotting

An equal amount of protein from cell lysates was mixed with 4x Laemmli buffer, subjected to SDS-PAGE, and transferred to polyvinylidene difluoride membranes. Equal loading was confirmed using MemCode protein stain (no. 24585, Thermo Fisher Scientific, Waltham, MA. Membranes were blocked with 5% nonfat milk (no.170-6404, Bio-Rad Laboratories, Hercules, CA) in TBST (Tris-buffered saline, pH 7.5 plus 0.1% Tween-20) for 1 h at room temperature, washed 3 times in TBST, and then incubated overnight with OXPHOS primary antibody cocktail (ab110411, Abcam, Cambridge, UK) at 1:1,000 dilution in 5% BSA (bovied serum albumin). Membranes were washed (3 times in TBST) and incubated at room temperature in 1:5,000 dilution of anti-mouse IgG horseradish peroxidase conjugate (no. 7076, Cell Signaling, Danvers, MA). Membranes were washed once more (3 times in TBST) and subjected to enhanced chemiluminescence and quantified by densitometry. Results are expressed relative to the normalized average of all the samples on the blot.

### 4.12. Statistical Analysis

The data is presented as ± S.E.M. Unless indicated otherwise, the data was analyzed using two-tailed Student’s paired t test for comparisons between timepoints. Comparisons between more than two timepoints were performed using either a one-way or two-way Analysis of Variance (ANOVA). Corrections for multiple comparisons were performed using the Benjamini-Hochberg procedure. All statistical analyses were performed using GraphPad Prism 10.3.1.

## AUTHORSHIP CONTRIBUTION STATEMENT

**Devanshi Gupta:** Investigation, Formal analysis, Data curation, Visualization, Writing – original draft, Writing – review & editing. **Kyle J. Sevits:** Investigation, Methodology, Formal analysis, Writing – review & editing. **Katherine A. Klaus:** Investigation, Methodology, Writing – review & editing. **Shiana S. Loona:** Investigation, Data curation, Writing – review & editing. **Vidhi D. Parmar:** Investigation, Data curation, Writing – review & editing. **Yohan Kim:** Investigation, Formal analysis, Writing – review & editing. **Carrie J. Heppelmann:** Investigation, Formal analysis, Writing – review & editing. **Robert G. Leija:** Investigation, Data curation, Writing – review & editing. **Hawley E. Kunz:** Formal analysis, Data interpretation, Writing – review & editing. **Fabrice Lucien:** Writing – review & editing. **Linda M. Szymanski:** Clinical oversight, Supervision, Writing – review & editing. **Aoife M. Egan:** Clinical oversight, Supervision, Writing – review & editing. **Mark W. Pataky:** Conceptualization, Methodology, Investigation, Supervision, Funding acquisition, Data curation, Writing – original draft, Writing – review & editing, Project administration.

## ACKNOWLEDGEMENTS

The authors thank Melissa Aakre, Rose Decker, Roberta Soderberg, Hannah Christie, and Talyia Fordham, as well as staff at the Clinical Research Unit at Mayo Clinic for their technical contributions. Special thanks are also given to Leanne Pataky, spouse of the corresponding author (M.W.P.), who raised the foundational question that inspired the work presented here, and to thank Mandi Trahan, Megan Spiten, Amanda Nygard, and Almira Smailovic for their assistance in preliminary studies.

## DATA AVAILABILITY

All data generated or analyzed during this study are included in this article and its supplementary Information files. Source data are provided. All raw data will be available upon acceptance to a peer reviewed journal. Any additional information required to reanalyze the data reported in this paper is available from the lead contact upon request.

## SUPPLEMENTAL INFORMATION

**Figure S1.**
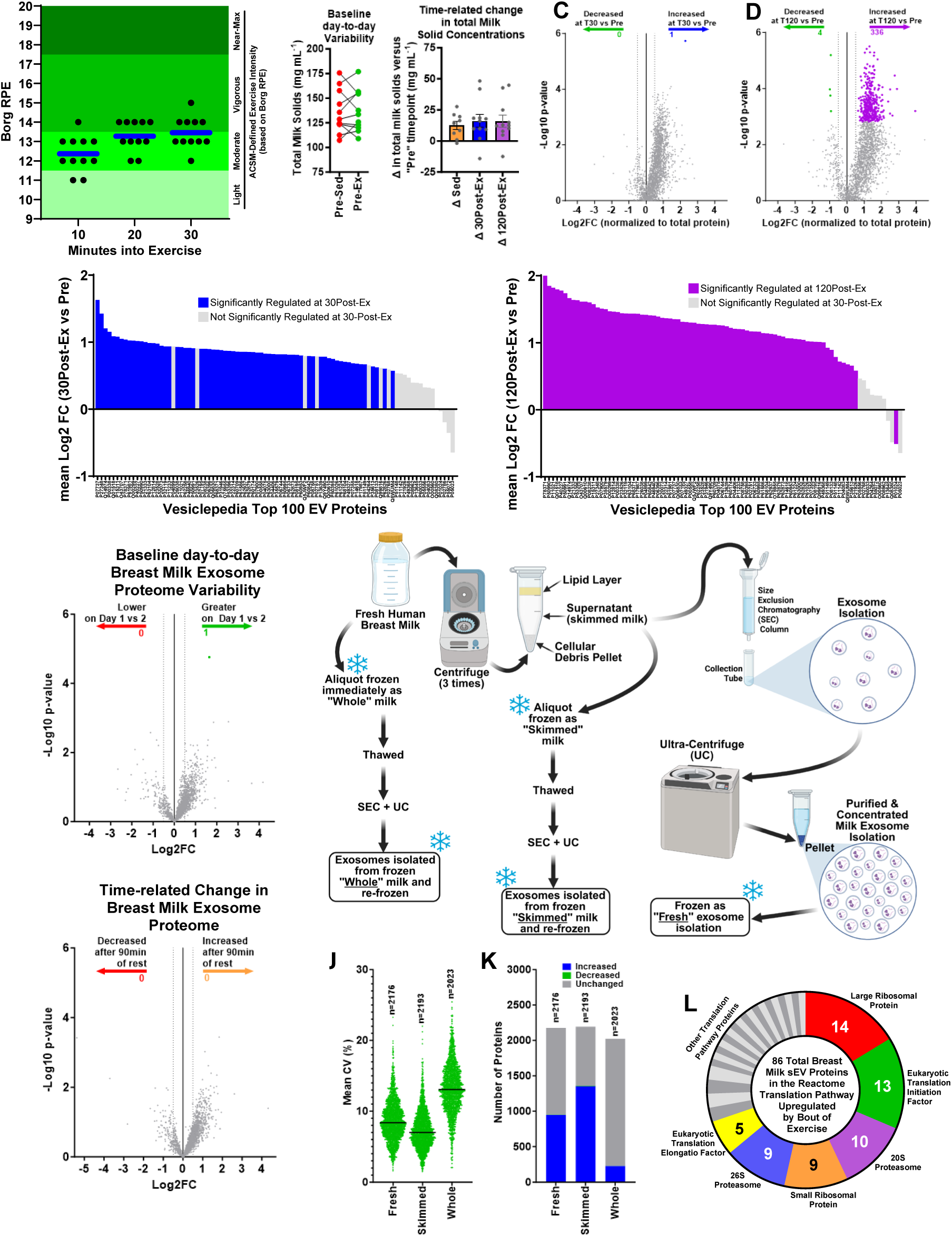
Methodological Validation Confirming Exercise-mediated Effects in Breast Milk sEVs. (**A**) Rating of perceived exertion (Borg) was assessed during exercise to confirm moderate-intensity. (**B**) Breast milk concentration across timepoints was measured by weighing milk solids from freeze dried whole milk. Data are presented as the mean ± SEM. A two-tailed paired t-test was used to compare day-to-day differences. Two-way ANOVA was used to compare time-related changes. No significant differences were observed. (left panel: *n*=11 per group; right panel: n=9 for Δ Sed, and n=11 for Δ PostEx comparisons). (**C and D**) Breast milk sEV proteomic changes at 30 (blue) and 120 (purple) minutes following a moderate-intensity exercise session versus pre-exercise (green) expressed relative tot total protein. Unadjusted p-value is plotted on the y-axis, but only proteins with FDR-corrected (Benjamini-Hochberg) q-value of < 0.1, and |log2FC| > 0.5 are colored (*n*=11 per group). (**E and F**) Post-exercise change in known EV database proteins. Proteins with FDR-corrected (Benjamini-Hochberg) q-value of < 0.1 and |log2FC| > 0.5 of are colored (*n*=11 per group). (**G and H**) Lack of breast milk sEV proteomic changes across study days at the baseline timepoint and following sedentary period. Unadjusted p-value is plotted on the y-axis, but only proteins with FDR-corrected (Benjamini-Hochberg) q-value of < 0.1 and |log2FC| > 0.5 are colored (one protein in panel e) (*n*=11 and *n*=9 in panels e and f, respectively). (**I**) Experimental procedures for isolation of breast milk sEVs from fresh and frozen samples. (**J**) Mean coefficient of variation for each identified protein (dots) from the “Pre-Ex” timepoint in sEVs isolated from fresh milk, frozen skimmed milk, and frozen whole milk (*n*=11). (**K**) Number of proteins that are unchanged (grey), significantly increased (blue) or decreased (green) at 30 minutes post-exercise from sEVs isolated from fresh milk, frozen skimmed milk, and frozen whole milk (*n*=11). Only proteins with FDR-corrected (Benjamini-Hochberg) q-value of < 0.1, and |log2FC| >0.5 are considered differentially expressed. (**L**) 86 breast milk sEV proteins in the Reactome Translation pathway were upregulated at both 30 and 120 minutes post-exercise, among which 70% were subunits of 6 proteins, highlighted in colored slices.

**Figure S2.**
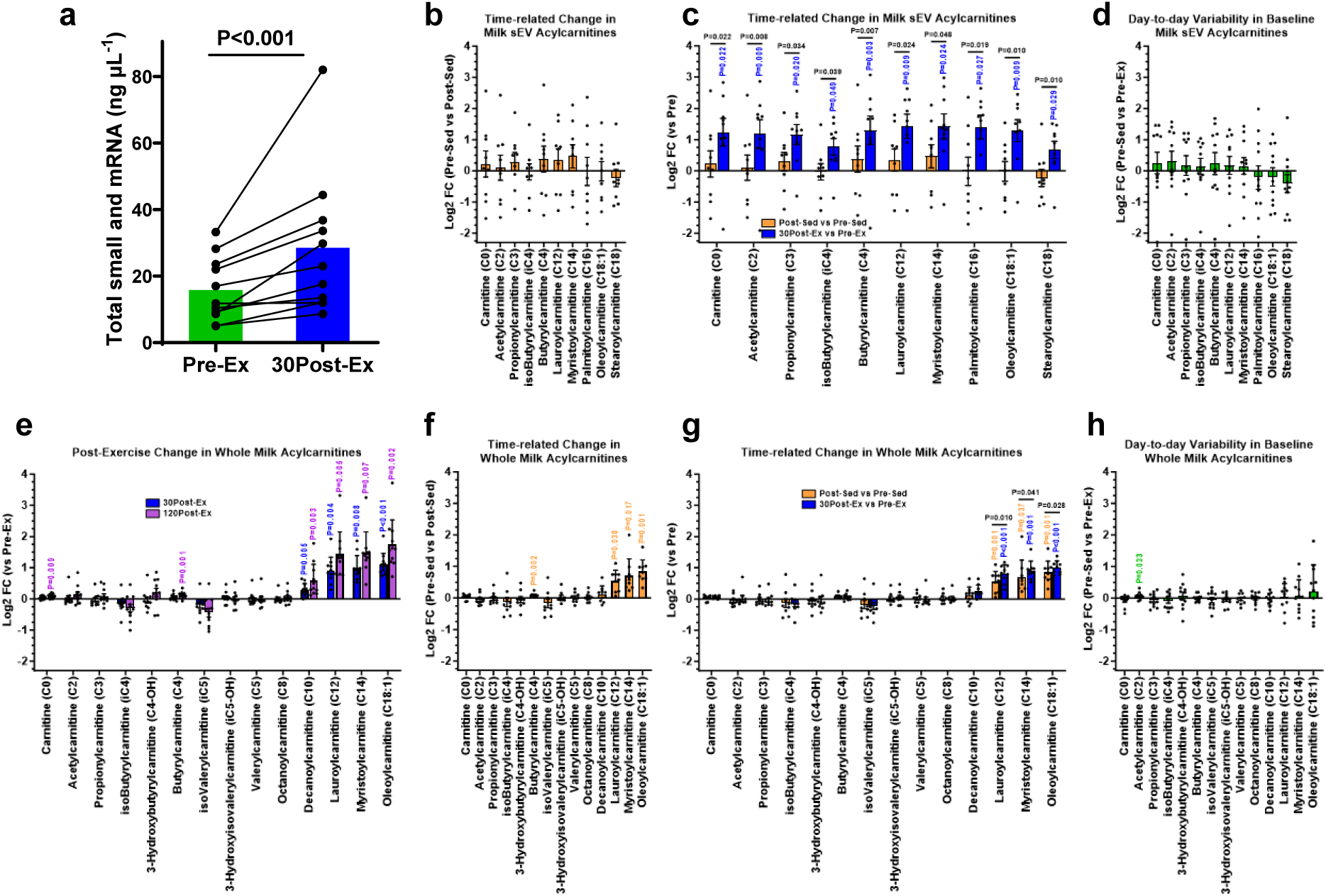
The Exercise Effect on Breast Milk sEVs is Not Due to a Time-related Change, as Demonstrated in Whole Milk. (**A**) Total small RNA and mRNA concentration from milk sEVs obtained before and 30 minutes after exercise. Lines represent individual participants (*n*=11). A two-tailed paired t-test was used to compare timepoints. (**B**) Change in breast milk sEV acylcarnitine species while sedentary. (**C**) Post-exercise change (blue bars) in breast milk sEV acylcarnitine species plotted on the same graph as post-sedentary changes (orange bars). Although pre- vs post-exercise data exists for 11 participants (see Fig. 2e), only participants with pre- vs post-sedentary data are presented for statistical comparisons (*n*=9 per group). (**D**) Baseline day-to-day variability in breast milk sEV acylcarnitine species is demonstrated by comparing Pre-Sed versus Pre-Ex timepoints. (**E**) Post-exercise changes in whole milk acylcarnitine species. (**F**) Post-sedentary changes in whole milk acylcarnitine species. (**G**) Post-exercise and post-sedentary changes in whole milk acylcarnitine species plotted on the same graph. Although pre- vs post-exercise data exists for 11 participants (see panel e), only participants with pre- vs post-sedentary data are presented for statistical comparisons (*n*=9 per group). (**H**) Baseline day-to-day variability in whole milk acylcarnitine species is demonstrated by comparing Pre-Sed versus Pre-Ex timepoints. The quantified data in (B)-(H) are presented as the mean ± SEM. Dots represent individual participants (*n*=9 for b, c, f, and g; n=11 for d, e, and h). For (B), (D), (F), and (H) two-tailed paired t-tests between groups (displayed on y axis) were used and P-values are shown on the graphs. For (C) and (G), two-way repeated measures ANOVAs were used to test the change in acylcarnitine concentrations on the sedentary versus exercise visit. P-values in black represent significant group x time interaction. P-values representing significant effects after multiple comparison testing in the Pre-Sed vs Post-Sed (orange) and the Pre-Ex vs 30Post-Ex (blue) are also shown. For (E), one-way ANOVA was used to test the time-related change in whole milk acylcarnitines on the exercise visit, and p values represent significant changes at 30Post-Ex (blue) and 120Post-Ex (purple) versus Pre-Ex.

**Figure S3.**
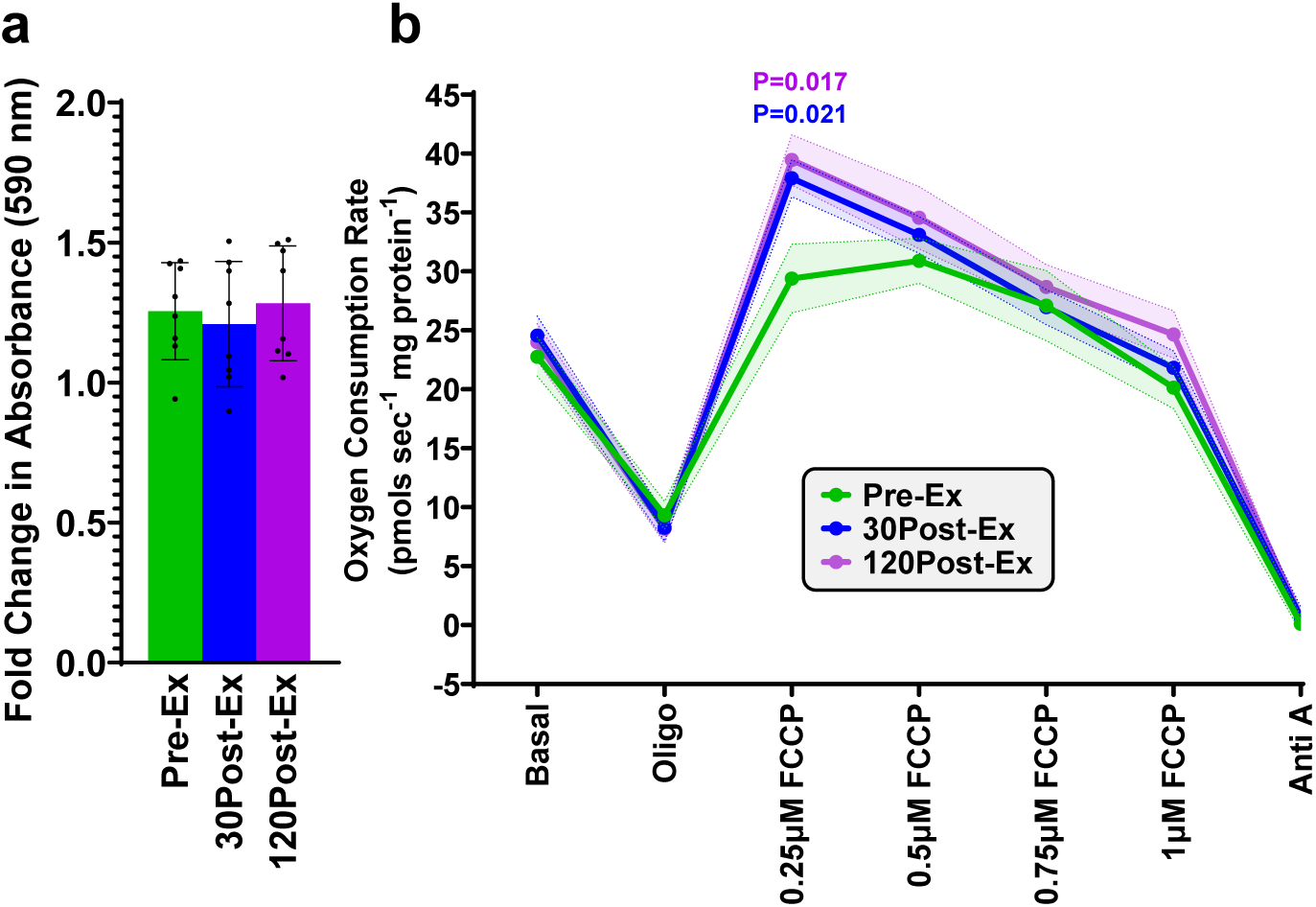
Exercised sEV Effects on UC-MSCs. (**A**) Quantitative analysis of solubilized dye was measured at 590nm absorbance. Fold change in absorbance from cells prior to exposure and 48hr after exposure or breast milk sEVs under three different conditions (Pre-Ex, 30Post-Ex, and 120Post-Ex) is displayed. Each sample was run in triplicate wells on a 96-well plate. Dots represent data from an individual sample. (**B**) Oxygen consumption rate curves of UC-MSCs exposed to breast milk sEVs isolated at Pre-Ex, 30Post-Ex, and 120Post-Ex in the basal state and in the presence of oligomycin (2 µg ml^−1^), sequential additions of 0.25 µM FCCP, and Antimycin A (2.5 µM). Statistical significance was calculated using one-way ANOVA followed by multiple comparisons (*n*=8 per group). P-values represent significant changes at 30Post-Ex (blue) and 120Post-Ex (purple) versus Pre-Ex at 0.25 µM FCCP. For quantified data in (A)-(B), data are presented as the mean ± SEM.

## REFERENCES

[1] De Boo, H.A., Harding, J.E., 2006. The developmental origins of adult disease (Barker) hypothesis. Australian and New Zealand Journal of Obstetrics and Gynaecology 46(1):4–14.

[2] Organization, W.H., 2000. Effect of breastfeeding on infant and child mortality due to infectious diseases in less developed countries: a pooled analysis. The Lancet 355(9197):451–455.

[3] Owen, C.G., Martin, R.M., Whincup, P.H., Smith, G.D., Cook, D.G., 2005. Effect of infant feeding on the risk of obesity across the life course: a quantitative review of published evidence. Pediatrics 115(5):1367–1377.

[4] Armstrong, J., Reilly, J.J., 2002. Breastfeeding and lowering the risk of childhood obesity. The Lancet 359(9322):2003–2004.

[5] Moholdt, T., Stanford, K.I., 2024. Exercised breastmilk: a kick-start to prevent childhood obesity? Trends in Endocrinology & Metabolism 35(1):23–30.

[6] Lovelady, C., Lonnerdal, B., Dewey, K.G., 1990. Lactation performance of exercising women. The American journal of clinical nutrition 52(1):103–109.

[7] Dewey, K.G., Lovelady, C.A., Nommsen-Rivers, L.A., McCrory, M.A., Lonnerdal, B., 1994. A randomized study of the effects of aerobic exercise by lactating women on breast-milk volume and composition. New England Journal of Medicine 330(7):449–453.

[8] Wright, K.S., Quinn, T.J., Carey, G.B., 2002. Infant acceptance of breast milk after maternal exercise. Pediatrics 109(4):585–589.

[9] Daley, A.J., Thomas, A., Cooper, H., Fitzpatrick, H., McDonald, C., Moore, H., et al., 2012. Maternal exercise and growth in breastfed infants: a meta-analysis of randomized controlled trials. Pediatrics 130(1):108–114.

[10] Harris, J.E., Pinckard, K.M., Wright, K.R., Baer, L.A., Arts, P.J., Abay, E., et al., 2020. Exercise-induced 3′-sialyllactose in breast milk is a critical mediator to improve metabolic health and cardiac function in mouse offspring. Nature metabolism 2(8):678–687.

[11] Admyre, C., Johansson, S.M., Qazi, K.R., Filén, J.-J., Lahesmaa, R., Norman, M., et al., 2007. Exosomes with immune modulatory features are present in human breast milk. The Journal of immunology 179(3):1969–1978.

[12] van Herwijnen, M.J., Zonneveld, M.I., Goerdayal, S., Nolte, E.N., Garssen, J., Stahl, B., et al., 2016. Comprehensive proteomic analysis of human milk-derived extracellular vesicles unveils a novel functional proteome distinct from other milk components. Molecular & Cellular Proteomics 15(11):3412–3423.

[13] Zhou, Q., Li, M., Wang, X., Li, Q., Wang, T., Zhu, Q., et al., 2012. Immune-related microRNAs are abundant in breast milk exosomes. International journal of biological sciences 8(1):118.

[14] Ye, S., Li, D., Jiang, C., Wang, X., Dong, M., Wei, W., 2025. Lipidomic Analysis of Extracellular Vesicles in Human Breast Milk and Comparison with Milk Polar Lipids Using Ultraperformance Supercritical Fluid Chromatography-Mass Spectrometry. J Agric Food Chem 73(17):10644–10653.

[15] Liao, Y., Du, X., Li, J., Lönnerdal, B., 2017. Human milk exosomes and their microRNAs survive digestion in vitro and are taken up by human intestinal cells. Molecular nutrition & food research 61(11):1700082.

[16] Betker, J.L., Angle, B.M., Graner, M.W., Anchordoquy, T.J., 2019. The potential of exosomes from cow milk for oral delivery. Journal of pharmaceutical sciences 108(4):1496–1505.

[17] Manca, S., Upadhyaya, B., Mutai, E., Desaulniers, A.T., Cederberg, R.A., White, B.R., et al., 2018. Milk exosomes are bioavailable and distinct microRNA cargos have unique tissue distribution patterns. Scientific reports 8(1):11321.

[18] Samuel, M., Fonseka, P., Sanwlani, R., Gangoda, L., Chee, S.H., Keerthikumar, S., et al., 2021. Oral administration of bovine milk-derived extracellular vesicles induces senescence in the primary tumor but accelerates cancer metastasis. Nature communications 12(1):3950.

[19] Dai, C., Xu, Q., Li, L., Liu, Y., Qu, S., 2024. Milk extracellular vesicles: natural nanoparticles for enhancing oral drug delivery against bacterial infections. ACS Biomaterials Science & Engineering 10(4):1988–2000.

[20] Warren, M.R., Zhang, C., Vedadghavami, A., Bokvist, K., Dhal, P.K., Bajpayee, A.G., 2021. Milk exosomes with enhanced mucus penetrability for oral delivery of siRNA. Biomaterials science 9(12):4260–4277.

[21] Vanderboom, P.M., Dasari, S., Ruegsegger, G.N., Pataky, M.W., Lucien, F., Heppelmann, C.J., et al., 2021. A size-exclusion-based approach for purifying extracellular vesicles from human plasma. Cell reports methods 1(3).

[22] Whitham, M., Parker, B.L., Friedrichsen, M., Hingst, J.R., Hjorth, M., Hughes, W.E., et al., 2018. Extracellular vesicles provide a means for tissue crosstalk during exercise. Cell metabolism 27(1):237–251. e234.

[23] Ashcroft, S.P., Stocks, B., Egan, B., Zierath, J.R., 2024. Exercise induces tissue-specific adaptations to enhance cardiometabolic health. Cell metabolism 36(2):278–300.

[24] Chow, L.S., Gerszten, R.E., Taylor, J.M., Pedersen, B.K., Van Praag, H., Trappe, S., et al., 2022. Exerkines in health, resilience and disease. Nature Reviews Endocrinology 18(5):273–289.

[25] Zonneveld, M.I., Brisson, A.R., van Herwijnen, M.J., Tan, S., van de Lest, C.H., Redegeld, F.A., et al., 2014. Recovery of extracellular vesicles from human breast milk is influenced by sample collection and vesicle isolation procedures. Journal of extracellular vesicles 3(1):24215.

[26] Galley, J.D., Besner, G.E., 2020. The therapeutic potential of breast milk-derived extracellular vesicles. Nutrients 12(3):745.

[27] Bozkurt, Z.B., Samur, G., 2026. Modulation of miRNAs in Breast Milk via Maternal Diet and Body Composition: a Comprehensive Review. Nutrition reviews:nuaf289.

[28] Meng, Z., Zhou, D., Lv, D., Gan, Q., Liao, Y., Peng, Z., et al., 2023. Human milk extracellular vesicles enhance muscle growth and physical performance of immature mice associating with Akt/mTOR/p70s6k signaling pathway. J Nanobiotechnology 21(1):304.

[29] Lu, C., Dreyfuss, J.M., Hua, T., Wolfs, D., Nagel, E.M., Peña, A., et al., 2025. Maternal Physical Activity and its Relationship to the Human Milk Metabolome and Infant Body Composition. The Journal of Clinical Endocrinology & Metabolism:dgaf296.

[30] Jevtovic, F., Zheng, D., Claiborne, A., Biagioni, E.M., Wisseman, B.L., Krassovskaia, P.M., et al., 2024. Effects of maternal exercise on infant mesenchymal stem cell mitochondrial function, insulin action, and body composition in infancy. Physiological Reports 12(9):e16028.

[31] Saunderson, S.C., Dunn, A.C., Crocker, P.R., McLellan, A.D., 2014. CD169 mediates the capture of exosomes in spleen and lymph node. Blood, The Journal of the American Society of Hematology 123(2):208–216.

[32] Santos, M.F., Randa, J., Tai, D., Vistoli, G., Schahaf, N.A., Vittorio, S., et al., 2025. Physical exercise increases binding of POMC to blood extracellular vesicles. Proceedings of the National Academy of Sciences 122(51):e2525044122.

[33] Frühbeis, C., Helmig, S., Tug, S., Simon, P., Krämer-Albers, E.-M., 2015. Physical exercise induces rapid release of small extracellular vesicles into the circulation. Journal of extracellular vesicles 4(1):28239.

[34] Munir, J., Sadri, M., Zempleni, J., 2025. Tsg101 knockout in the mammary gland leads to a decrease in small extracellular vesicles in milk from C57BL/6J dams and contributes to leakiness of the gut mucosa and reduced postnatal weight gain in suckling pups. The Journal of Nutritional Biochemistry 135:109782.

[35] Cartee, G.D., 2015. Mechanisms for greater insulin-stimulated glucose uptake in normal and insulin-resistant skeletal muscle after acute exercise. American Journal of Physiology-Endocrinology and Metabolism 309(12):E949–E959.

[36] Lahti-Pulkkinen, M., Bhattacharya, S., Wild, S.H., Lindsay, R.S., Räikkönen, K., Norman, J.E., et al., 2019. Consequences of being overweight or obese during pregnancy on diabetes in the offspring: a record linkage study in Aberdeen, Scotland. Diabetologia 62(8):1412–1419.

[37] Carter, L.G., Lewis, K.N., Wilkerson, D.C., Tobia, C.M., Ngo Tenlep, S.Y., Shridas, P., et al., 2012. Perinatal exercise improves glucose homeostasis in adult offspring. American Journal of Physiology-Endocrinology and Metabolism 303(8):E1061–E1068.

[38] Laker, R.C., Altıntaş, A., Lillard, T.S., Zhang, M., Connelly, J.J., Sabik, O.L., et al., 2021. Exercise during pregnancy mitigates negative effects of parental obesity on metabolic function in adult mouse offspring. Journal of applied physiology 130(3):605–616.

[39] Stanford, K.I., Lee, M.-Y., Getchell, K.M., So, K., Hirshman, M.F., Goodyear, L.J., 2015. Exercise before and during pregnancy prevents the deleterious effects of maternal high-fat feeding on metabolic health of male offspring. Diabetes 64(2):427–433.

[40] Kusuyama, J., Alves-Wagner, A.B., Conlin, R.H., Makarewicz, N.S., Albertson, B.G., Prince, N.B., et al., 2021. Placental superoxide dismutase 3 mediates benefits of maternal exercise on offspring health. Cell metabolism 33(5):939–956. e938.

[41] Davenport, M.H., Meah, V.L., Ruchat, S.M., Davies, G.A., Skow, R.J., Barrowman, N., et al., 2018. Impact of prenatal exercise on neonatal and childhood outcomes: a systematic review and meta-analysis. Br J Sports Med 52(21):1386–1396.

[42] Chen, Y., Ma, G., Hu, Y., Yang, Q., Deavila, J.M., Zhu, M.J., et al., 2021. Effects of Maternal Exercise During Pregnancy on Perinatal Growth and Childhood Obesity Outcomes: A Meta-analysis and Meta-regression. Sports Med 51(11):2329–2347.

[43] Jevtovic, F., Collier, D.N., DeVente, J., Mouro, S., Claiborne, A., Wisseman, B., et al., 2024. Maternal exercise increases infant resting energy expenditure: preliminary results. International journal of obesity 48(9):1347–1350.

[44] Jevtovic, F., Claiborne, A., DeVente, J.E., Mouro, S., Houmard, J.A., Broskey, N.T., et al., 2025. Maternal resistance exercise increases infant energy expenditure. American Journal of Physiology-Endocrinology and Metabolism 328(3):E354–E361.

[45] Kyere-Davies, G., Hill, K.B., Mullen, G.P., Varshney, R.R., Das, S., Martinez, A., et al., 2026. Maternal exercise during lactation remodels obesity-associated mammary metabolism and milk fatty acids, enhancing offspring lipid oxidation. Am J Physiol Endocrinol Metab.

[46] Matsuda, A., Moirangthem, A., Angom, R.S., Ishiguro, K., Driscoll, J., Yan, I.K., et al., 2020. Safety of bovine milk derived extracellular vesicles used for delivery of RNA therapeutics in zebrafish and mice. Journal of Applied Toxicology 40(5):706–718.

[47] Holmen, M., Giskeødegård, G.F., Moholdt, T., 2023. High-intensity exercise increases breast milk adiponectin concentrations: a randomised cross-over study. Frontiers in nutrition 10:1275508.

[48] Holm, R.L., Holmen, M., Sujan, M.A.J., Giskeødegård, G.F., Moholdt, T., 2025. Acute effect of endurance exercise on human milk insulin concentrations: a randomised cross-over study. Frontiers in nutrition 11:1507156.

[49] Miclat, N.N., Hodgkinson, R., Marx, G.F., 1978. Neonatal gastric pH. Anesth Analg 57(1):98–101.

[50] Yu, G., Zheng, Q.-S., Li, G.-F., 2014. Similarities and differences in gastrointestinal physiology between neonates and adults: a physiologically based pharmacokinetic modeling perspective. The AAPS journal 16(6):1162–1166.

[51] Engfer, M.B., Stahl, B., Finke, B., Sawatzki, G., Daniel, H., 2000. Human milk oligosaccharides are resistant to enzymatic hydrolysis in the upper gastrointestinal tract. The American journal of clinical nutrition 71(6):1589–1596.

[52] Lemoine, M.C.C., Klau, L.J., Holmen, M., Ashby, E.R., Aachmann, F.L., Andreassen, T., et al., 2025. Unaltered 3’-sialyllactose and 6’-sialyllactose concentrations in human milk acutely after endurance exercise: a randomized crossover trial. Frontiers in nutrition 12:1638430.

[53] Wolfs, D., Lynes, M.D., Tseng, Y.-H., Pierce, S., Bussberg, V., Darkwah, A., et al., 2021. Brown fat–activating lipokine 12, 13-diHOME in human milk is associated with infant adiposity. The Journal of Clinical Endocrinology & Metabolism 106(2):e943–e956.

[54] Müller, M.J., Bosy-Westphal, A., Klaus, S., Kreymann, G., Lührmann, P.M., Neuhäuser-Berthold, M., et al., 2004. World Health Organization equations have shortcomings for predicting resting energy expenditure in persons from a modern, affluent population: generation of a new reference standard from a retrospective analysis of a German database of resting energy expenditure. The American journal of clinical nutrition 80(5):1379–1390.

[55] Bzikowska-Jura, A., Szulińska, A., Szostak-Węgierek, D., 2020. Resting energy expenditure during breastfeeding: body composition analysis vs. predictive equations based on anthropometric parameters. Nutrients 12(5):1274.

[56] Cimorelli, M., Nieuwland, R., Varga, Z., van der Pol, E., 2021. Standardized procedure to measure the size distribution of extracellular vesicles together with other particles in biofluids with microfluidic resistive pulse sensing. PLoS One 16(4):e0249603.

[57] Northrop-Albrecht, E.J., Kim, Y., Taylor, W.R., Majumder, S., Kisiel, J.B., Lucien, F., 2025. The proteomic landscape of stool-derived extracellular vesicles in patients with pre-cancerous lesions and colorectal cancer. Communications biology 8(1):228.

[58] Orsburn, B.C., 2021. Proteome discoverer—a community enhanced data processing suite for protein informatics. Proteomes 9(1):15.

[59] Sherman, B.T., Hao, M., Qiu, J., Jiao, X., Baseler, M.W., Lane, H.C., et al., 2022. DAVID: a web server for functional enrichment analysis and functional annotation of gene lists (2021 update). Nucleic acids research 50(W1):W216–W221.

[60] Wilkins, J., Sakrikar, D., Petterson, X.-M., Lanza, I.R., Trushina, E., 2019. A comprehensive protocol for multiplatform metabolomics analysis in patient-derived skin fibroblasts. Metabolomics 15(6):1–9.

[61] Pataky, M.W., Kumar, A.P., Gaul, D.A., Moore, S.G., Dasari, S., Robinson, M.M., et al., 2023. Divergent skeletal muscle metabolomic signatures of different exercise training modes independently predict cardiometabolic risk factors. Diabetes 73(1):23–37.

[62] Hagey, D.W., Ojansivu, M., Bostancioglu, B.R., Saher, O., Bost, J.P., Gustafsson, M.O., et al., 2023. The cellular response to extracellular vesicles is dependent on their cell source and dose. Science advances 9(35):eadh1168.

[63] Janssen, R.C., Boyle, K.E., 2019. Microplate assays for spectrophotometric measurement of mitochondrial enzyme activity. High-Throughput Metabolomics: Methods and Protocols. Springer, p. 355–368.

